# Impact of ventriculo-cisternal irrigation in preventing delayed cerebral ischemia in aneurysmal subarachnoid hemorrhage

**DOI:** 10.1101/2023.08.06.23293733

**Authors:** Motoyuki Umekawa, Gakushi Yoshikawa

## Abstract

**Background:** Delayed cerebral ischemia (DCI) due to vasospasm following subarachnoid hemorrhage (SAH) is considered a significant determinant of morbidity and mortality; however, no established method exists to prevent and treat vasospasm or DCI. This study aimed to evaluate the effectiveness of ventriculo-cisternal irrigation (VCI) in preventing vasospasms and DCI.

**Methods:** We retrospectively identified 340 SAH patients with ruptured intracranial aneurysms treated with postoperative VCI at our institution between December 2010 and January 2020. Ventricular/cisternal drainage (VD/CD) was inducted during aneurysm surgery, and lactated Ringer’s solution was used for irrigation until day 4 of SAH, followed by ICP control at 5–10 cmH_2_O until day 14. We collected data on total vasospasm, DCI, and modified Rankin Scale scores at discharge and analyzed the risk factors using logistic regression models.

**Results:** The median age was 65 years (interquartile range: 52–75), with 236 female patients (69%). The World Federation of Neurosurgical Societies grade distribution was as follows: Grade I or II, 175 cases (51%); Grade III or IV, 84 (25%); Grade V, 81 (24%). With VCI management in all cases, total vasospasm occurred in 162 patients (48%), but DCI incidence was low (23 patients [6.8%]). Major drainage-related complications were observed in five patients (1.5%). Early surgery, performed on SAH day 0 or 1, was identified as a preventive factor against DCI occurrence (odds ratio [OR] 0.21, 95% confidence interval [CI] 0.07–0.67; *p* = 0.008), while additional surgery (OR 4.76, 95% CI 1.62–13.98; *p* = 0.005) and dyslipidemia (OR 3.27, 95% CI 1.24–8.63; *p* = 0.017) were associated with DCI occurrence.

**Conclusions:** Managing vasospasms with VCI after SAH achieved a low incidence of 6.8% for DCI and is considered a safe and effective method. Early surgery after SAH occurrence was associated with a decreased risk of DCI with VCI therapy.

## Introduction

Subarachnoid hemorrhage (SAH) is one of the most severe cerebrovascular diseases, with 1– 20 patients per 100,000 incidence rates and approximately 85% of cases caused by rupture of intracranial aneurysms.^1, 2^ Its morbidity and mortality remain high, and re-ruptured aneurysms lead to approximately 50–60% mortality.^3, 4^ Although these conditions often inevitably occur at the onset of SAH, appropriate intensive interventions should be made to prevent re-rupture, surgery, or endovascular treatment.

Cerebral vasospasm followed by delayed cerebral ischemia (DCI) is the most difficult condition in the acute to subacute phase in patients with SAH and rarely results in severe neurological impairment. Its incidence rates have been reported as up to 70% for angiographic vasospasm and 20–40% for DCIs.^5–12^ However, the mechanism of vasospasms and standard preventative or efficacious treatments for vasospasms are not fully understood and established. We have treated many patients with aneurysmal SAH (including dissecting aneurysms) at our tertiary center and have adopted ventriculo-cisternal irrigation therapy (VCI) followed by intracranial pressure (ICP) control management as a unified method for preventing and treating vasospasms for over 20 years. This study aimed to present the outcomes of patients treated at our institution and evaluate the efficacy of VCI and ICP control in preventing vasospasms and DCIs.

## Methods

### Patient selection

Between December 2010 and January 2020, we retrospectively analyzed 340 consecutive patients who developed SAH due to ruptured cerebral aneurysms (including dissecting aneurysms) and underwent VCI therapy in our hospital. We collected data on the patient, aneurysm, SAH, and surgical factors. Informed consent was obtained from all participants and this study was approved by the Institutional Ethics Committee of the Showa General Hospital (approval number REC-328).

### Our principles for postoperative management of SAH

The treatment algorithm for aneurysmal SAH is presented in **Figure 1A**. When performing clipping or trapping surgery for SAH in Fisher groups 2 or 3, we performed VCI followed by 2 weeks of ICP control to prevent vasospasm. During surgery, we intraoperatively placed a ventricular drainage (VD) to manage cerebrospinal fluid (CSF) drainage and postoperative irrigation or drainage. Additionally, we placed a cisternal drainage (CD) or lumbar drainage (LD) for the CSF drainage system, which was used for postoperative management.

**Figure 1.**
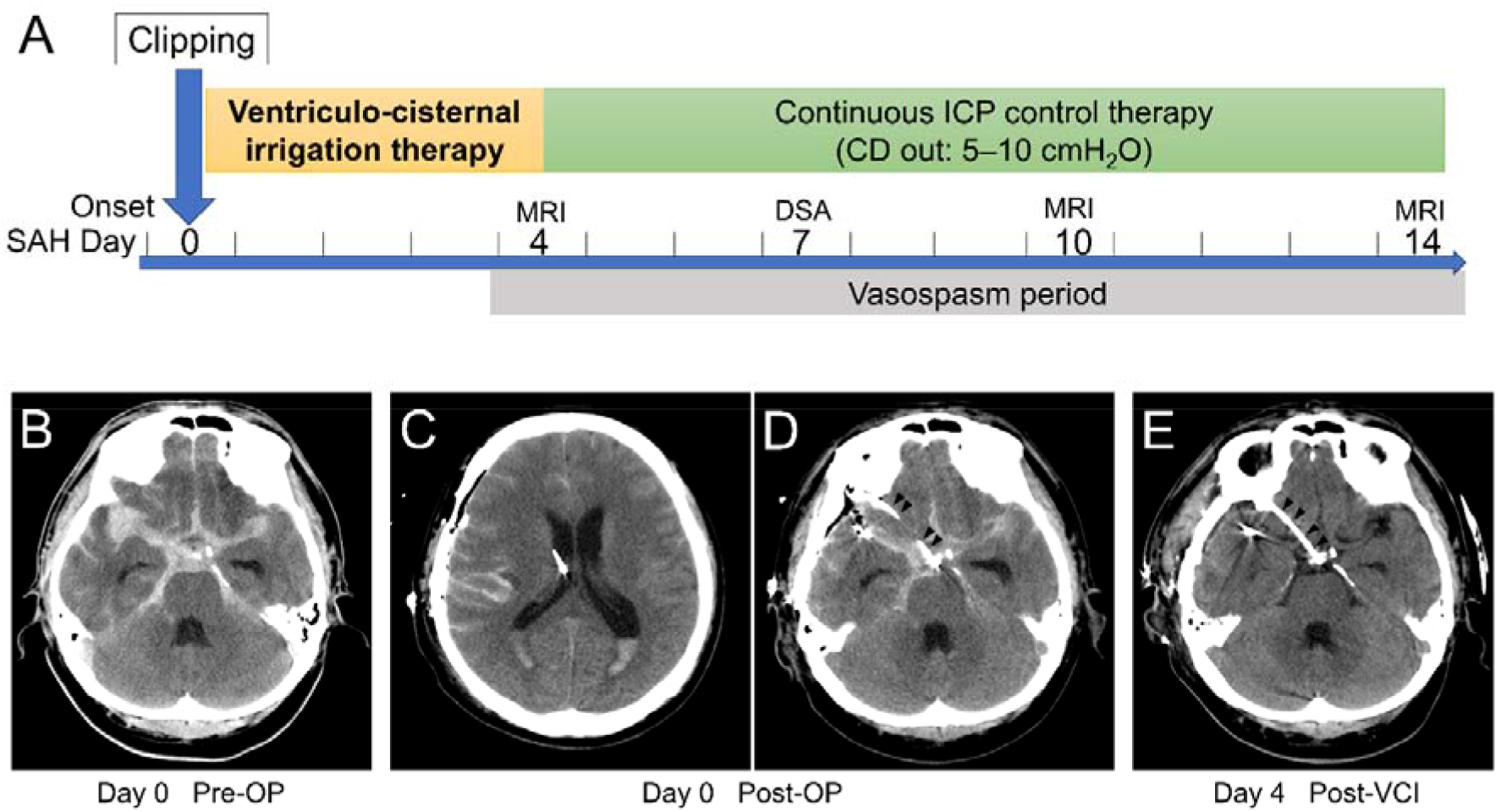
Scheme of our principles of surgical treatment in acute phase for aneurysmal subarachnoid hemorrhage (SAH) with ventriculo-cisternal irrigation (VCI) and continuous intracranial pressure (ICP) control therapy (**A**). Our approach to vasospasm prevention includes: 1) maximum washout of SAH by VCI until SAH Day4, 2) continuous drainage with CD for ICP control during the vasospasm period after Day 4 to 14, and 3) intensive care management and rehabilitation. Demonstrative case with SAH with ruptured right middle cerebral artery aneurysm (initial CT image; **B**) were treated with our standardized method. On the day of SAH onset, clipping of aneurysm was performed and ventricular drainage (**C**) and cisternal drainage (indicated by arrowheads; **D**) were placed. After four days of VCI therapy, SAH was completely washed out (**E**). CD, cisternal drainage; CT, computed tomography; DSA, digital subtraction angiography; ICP, intracranial pressure; OP, operation; SAH, subarachnoid hemorrhage; VD, ventricular drainage

We administered a drop infusion system for irrigation with lactated Ringer’s solution via the VD at a rate of 20 mL/h and released from the CD at the height of the forehead in the supine position. Starting on postoperative day 1, urokinase was added to the lactated Ringer’s solution at a concentration of 120 U/mL to expedite the removal of clots through VCI. In cases involving intraventricular hemorrhage, we initiated VCI inversely with drainage from the CD and infusion from the VD. Once the intraventricular hematoma had resolved, we switched to regular circulation with VD infusion and CD drainage. After confirmation of SAH washout from computed tomography (CT) images taken on day 4, we extracted the VD tube and completed the VCI. Thereafter, we continued CSF drainage with CD for ICP control around SAH day 14; the CD was opened at the height of the forehead with the patient supine at a head-up position of 20°−30°. Representative images are shown in **Figure 1B-E**. Cefazolin was administered intravenously during drainage as prophylaxis against meningitis.

Regarding imaging evaluation, we primarily assessed the distribution of SAH using CT scans until day 3. On day 4, magnetic resonance imaging (MRI) was performed to assess the early ischemic lesions and vascular conditions during the pre-vasospasm phase. On day 7, we performed digital subtraction angiography to evaluate the presence of cerebral vasospasm and aneurysms. We performed MRI to detect the presence of late-stage vasospasms and ischemic lesions on day 14.

### Details of ventriculo-cisternal irrigation and evaluated outcomes

We conducted a detailed investigation on VCI therapy, including the route and duration, presence of urokinase, and ICP control period during VCI therapy. The primary outcomes were the incidence of vasospasm/DCI and the symptoms caused by DCI. The secondary outcome was the modified Rankin Scale (mRS) score at discharge. Additionally, we conducted a detailed safety assessment of the complications related to drainage management.

### Statistical analysis

The median and interquartile ranges (IQR) were calculated for each factor. Bivariate and multivariate logistic regression analyses were performed to determine the occurrence of vasospasms and DCI for each factor. The favorable outcome group at discharge was defined as an mRS score of 0–2, and the same analysis was conducted for this group. A *p*-value < 0.05 was considered statistically significant. The factors used in the multivariate analysis were selected using a stepwise forward selection method with a *p*-value threshold of less than 0.15. Statistical analyses were performed using JMP Pro 17 software (SAS Institute Inc., Cary, NC, USA).

## Results

### Characteristics of patients, SAH, and surgeries

The patient demographics and SAH factors are shown in **Table 1**. The median age was 65 years (IQR, 52–75 years), with female patients accounting for 236 cases (69%). Hypertension was the most common comorbidity and risk factor for arterial atherosclerosis. The World Federation of Neurosurgical Societies (WFNS) grades were as follows: grade I, 72 patients (21%); II, 103 (30%); III, eight (2%); IV, 76 (22%); and V, 81 (24%). In the entire cohort, saccular aneurysms accounted for the majority, with 319 cases (94%), while 307 cases (90%) were treated for anterior circulation aneurysms. The most common location of aneurysms was the internal carotid artery-posterior communicating artery, with 88 cases (26%), followed by the anterior communicating artery, with 86 cases (25%), and the middle cerebral artery, with 74 cases (22%).

**Table 1.** Baseline characteristics of patients with aneurysmal subarachnoid hemorrhage.

|  | Number (%) / Median [IQR] |
| --- | --- |
| Age, years | 65 [52–75] |
| Female sex | 236 (69%) |
| Comorbidities |  |
| Hypertension | 190 (56%) |
| Dyslipidemia | 62 (18%) |
| Diabetes mellitus | 22 (6%) |
| Active smoking | 93 (27%) |
| Familial history of intracranial aneurysm | 31 (9%) |
| Use of antithrombotic | 19 (6%) |
| WFNS grading |  |
| I | 72 (21%) |
| II | 103 (30%) |
| III | 8 (2%) |
| IV | 76 (22%) |
| V | 81 (24%) |
| Aneurysm type |  |
| Saccular | 319 (94%) |
| Maximum dome diameter, mm | 5.1 [4.0–7.6] |
| Dissecting | 21 (6%) |
| Maximum length, mm | 5.4 [3.0–9.3] |
| Location of aneurysm |  |
| IC-Pcom | 88 (26%) |
| Acom | 86 (25%) |
| MCA | 74 (22%) |
| Distal ACA | 21 (6%) |
| IC-Ach | 20 (6%) |
| Other anterior circulation | 18 (5%) |
| VA | 14 (4%) |
| BA | 12 (4%) |
| Other posterior circulation | 7 (2%) |
| Concomitant intracerebral hemorrhage | 83 (24%) |
| Preoperative anisocoria/dilated pupils | 45 (13%) |

ACA, anterior cerebral artery; Acom, anterior communicating artery; BA, basilar artery; IC-Pcom, internal carotid artery-posterior communicating artery; IQR, interquartile range; MCA, middle cerebral artery; VA, vertebral artery; WFNS, The World Federation of Neurosurgery

The surgical methods used were aneurysmal neck clipping in 323 patients (95%) and parental artery trapping or proximal occlusion in 17 patients (5%) (**Supplementary Table 1**). Additional techniques included bypass in 23 patients (7%) and combined decompressive craniectomy in 22 patients (6%). VD before aneurysm treatment was performed in 44 patients (13%). The day of surgery for aneurysm treatment was day 0 (onset of SAH) in 193 patients (57%) and day 1 in 116 patients (34%), while treatment on day 2 or later was performed in 31 patients (9%). Early surgery on day 0 or 1 was performed more frequently in the WFNS grade IV/V group (98.1% vs. 84.7% in grades I–III group; *p* <0.001). Additional surgery, including hematoma evacuation and decompressive craniectomy, was required in 31 patients (9%).

### Details of ventriculo-cisternal irrigation and ICP control therapy

VCI was performed in 268 patients (79%) using VD-CD irrigation and 72 patients (21%) using VD-LD irrigation. Urokinase solution was used with VCI in 244 patients (72%). The median duration of VCI was 5 days (IQR, 4–6 days), followed by a median duration of 26 days (IQR, 14–16 days) for ICP control with CD or LD (**Supplementary Table 2**).

All complications associated with drainage management were observed in 22 cases (6.5%), including five cases (1.5%) of severe complications resulting in permanent disability or requiring surgical intervention. The most common complication was bleeding around the ventricular drain in 10 cases (2.9%), of which 2 cases (0.6%) required additional hematoma evacuation, while the rest had minor bleeding without residual symptoms. Although the syndrome of intracranial hypotension during ICP management was observed in six cases (1.8%), only one case (0.3%) required treatment, which involved hematoma evacuation for a surgical epidural hematoma. Eventually, the patient fully recovered with a favorable outcome without any impairment (**Supplementary Table 3**).

### Cerebral vasospasm and delayed cerebral ischemia: incidence and risk analysis

Of the 340 cases, 162 (47%) exhibited total vasospasms (**Table 2**). Of these, 139 patients (41%) had transient vasospasms, and 23 (6.8%) eventually developed into DCI. Among the 23 patients with DCI, the most common symptom was hemiparalysis in 15 (4.4%), followed by disturbed consciousness in 8 (2.3%), aphasia in 7 (2.1%), other impairments of cognitive function in 5 (1.5%), and sensory disturbances in 2 (0.6%). The group that underwent early surgery (days 0, 1) had a DCI incidence of 5.8%, whereas the group that underwent surgery after day 2 had an incidence of 16.1% (chi-square, *p* = 0.029).

**Table 2.** Cerebral vasospasm and delayed cerebral ischemia after aneurysmal subarachnoid hemorrhage treated with surgery.

|  | Number (%) |
| --- | --- |
| Total vasospasm | 162 (47.6%) |
| Transient vasospasm | 139 (40.9%) |
| Delayed cerebral infarction | 23 (6.8%) |
| Motor paresis | 15 (4.4%) |
| Consciousness disturbance | 8 (2.3%) |
| Aphasia | 7 (2.1%) |
| Other cognitive function disturbance | 5 (1.5%) |
| Sensory disturbance | 2 (0.6%) |

The results of the nominal logistic analysis of the risk factors associated with total vasospasm and DCI occurrence are shown in **Tables 3 and 4**. In the bivariate analysis of total vasospasm, statistically significant risk factors were female, anterior circulation aneurysm, and using the pterional approach. In the multivariate analysis, female (OR 1.64; 95% CI 1.01–2.67; *p* = 0.047) and anterior circulation aneurysm (OR 5.17; 95% CI 1.93–13.83; *p* = 0.001) remained statistically significant risk factors. Early surgery is borderline for a significantly lower risk of vasospasm (OR 0.47, 95% CI 0.22–1.04; *p* = 0.064). In the bivariate analysis of DCI occurrence, related risk factors were dyslipidemia and postoperative additional surgery, while early surgery after SAH onset significantly reduced the risk of DCI. The multivariate analysis also identified dyslipidemia (OR 3.27; 95% CI 1.24–8.63; *p* = 0.017) and additional postoperative surgery (OR 4.76; 95% CI 1.62–13.98; *p* = 0.005) as statistically significant risk factors for DCI, with early surgery after SAH onset similarly reducing the risk of DCI (OR 0.21; 95% CI 0.07–0.67; *p* = 0.008).

**Table 3.** Risk analyses related to total vasospasms after SAH surgery under ventriculo-cisternal irrigation.

|  | Bivariate |  | Multivariate |
| --- | --- | --- | --- |

|  | OR [95% CI] | *p*-value | OR [95% CI] | *p*-value |
| --- | --- | --- | --- | --- |
| <u>Patient factor</u> |  |  |  |  |
| Age, years (continuous) | 1.00 [0.98–1.01] | 0.548 |  |  |
| Age ≥65, years | 0.93 [0.61–1.43] | 0.749 |  |  |
| **Female sex** | **1.62 [1.01–2.58]** | **0.045\*** | **1.64 [1.01–2.67]** | **0.047\*** |
| Hypertension | 1.36 [0.89–2.10] | 0.158 |  |  |
| Dyslipidemia | 1.04 [0.60–1.80] | 0.897 |  |  |
| Diabetes mellitus | 0.91 [0.38–2.17] | 0.831 |  |  |
| Active smoking | 0.69 [0.42–1.11] | 0.125 |  |  |
| Familial history of cerebral aneurysm | 1.37 [0.65–2.88] | 0.402 |  |  |
| Use of antithrombotic | 0.49 [0.18–1.32] | 0.156 |  |  |
| <u>SAH factor</u> |  |  |  |  |
| Dissection (vs. saccular) | 0.81 [0.33–1.98] | 0.651 |  |  |
| **Anterior circulation (vs. posterior circulation)** | **5.86 [2.21–15.58]** | **<0.001\*** | **5.17 [1.93–13.83]** | **0.001\*** |
| With ICH (vs. without ICH) | 1.51 [0.92–2.49] | 0.104 |  |  |
| WFNS grade 4, 5 (vs. grade 1–3) | 0.88 [0.57–1.34] | 0.541 |  |  |
| <u>Operative factor</u> |  |  |  |  |
| PA (vs. other approaches) | **1.89 [1.15–3.15]** | **0.012\*** |  |  |
| Clipping (vs. others) | 1.71 [0.62–4.74] | 0.301 |  |  |
| Bypass | 1.21 [0.52–2.88] | 0.653 |  |  |
| Decompressive craniotomy | 2.01 [0.82–4.93] | 0.127 | 2.06 [0.82–5.14] | 0.122 |
| Emergent ventricular drainage | 0.90 [0.48–1.71] | 0.755 |  |  |
| Early surgery at day 0, 1 (vs. delayed surgery after day 2) | 0.47 [0.22–1.01] | 0.053 | 0.47 [0.22–1.04] | 0.064 |
| Postoperative rerupture | 1.80 [0.58–5.61] | 0.313 |  |  |
| Postoperative additional surgery | 1.84 [0.86–3.92] | 0.115 |  |  |

CI, confidence interval; ICH, intracerebral hemorrhage; OR, odds ratio; PA, pterional approach; WFNS, World Federation of Neurosurgical Societies

**Table 4.** Risk analyses related to delayed cerebral infarction after SAH surgery under ventriculo-cisternal irrigation.

|  | Bivariate |  | Multivariate |  |
| --- | --- | --- | --- | --- |
|  | OR [95% CI] | *p*-value | OR [95% CI] | *p*-value |
| <u>Patient factor</u> |  |  |  |  |
| Age, years (continuous) | 1.01 [0.98–1.04] | 0.560 |  |  |
| Age ≥65, years | 1.31 [0.56–3.07] | 0.537 |  |  |
| Female sex | 2.19 [0.73–6.60] | 0.164 |  |  |
| Hypertension | 1.25 [0.52–2.96] | 0.619 |  |  |
| **Dyslipidemia** | **2.60 [1.05–6.43]** | **0.039\*** | **3.27 [1.24–8.63]** | **0.017\*** |
| Diabetes mellitus | 1.41 [0.31–6.46] | 0.655 |  |  |
| Active smoking | 0.24 [0.05–1.03] | 0.055 | 0.24 [0.05–1.09] | 0.064 |
| Familial history of cerebral aneurysm | 0.95 [0.21–4.24] | 0.942 |  |  |
| Use of antithrombotic | 2.82 [0.76–10.50] | 0.122 |  |  |
| <u>SAH factor</u> |  |  |  |  |
| Dissection (vs. saccular) | 2.5 [0.68–9.17] | 0.170 |  |  |
| Anterior circulation (vs. posterior circulation) | 1.14 [0.25–5.09] | 0.866 |  |  |
| With ICH (vs. without ICH) | 1.39 [0.55–3.50] | 0.488 |  |  |
| WFNS grade 4, 5 (vs. grade 1–3) | 0.73 [0.31–1.75] | 0.484 |  |  |
| <u>Operative factor</u> |  |  |  |  |
| PA (vs. other approaches) | 1.24 [0.44–3.43] | 0.685 |  |  |
| Clipping (vs. others) | 0.52 [0.11–2.43] | 0.408 |  |  |
| Bypass | 2.23 [0.61–8.13] | 0.226 |  |  |
| Decompressive craniotomy | 0.64 [0.08–4.99] | 0.671 |  |  |
| Emergent ventricular drainage | 1.46 [0.47–4.5] | 0.512 |  |  |
| **Early surgery at day 0, 1 (vs. delayed surgery after day 2)** | **0.32 [0.11–0.94]** | **0.038\*** | **0.21 [0.07–0.67]** | **0.008\*** |
| Postoperative rerupture | 2.65 [0.55–12.74] | 0.224 |  |  |
| **Postoperative additional surgery** | **4.12 [1.49–11.39]** | **0.006\*** | **4.76 [1.62–13.98]** | **0.005\*** |

CI, confidence interval; ICH, intracerebral hemorrhage; OR, odds ratio; PA, pterional approach; WFNS, World Federation of Neurosurgical Societies

### Overall clinical outcomes

At discharge, the outcomes of performance status were as follows: a favorable outcome (mRS 0–2) was observed in 151 cases (44%), moderate outcome (mRS 3, 4) in 125 cases (37%), and severe outcome (mRS 5, 6) in 64 cases (19%) (**Supplementary Table 4**). In the entire cohort, the mortality rate was 5.3%. Stratified by the WFNS grade, treatment of grades I and II achieved 79% and 64% of favorable outcomes at discharge (**Figure 2**). In the grade IV and V groups, although 21% and 52%, respectively, ended up with severe outcomes (mRS 5, 6) at discharge, intermediate outcomes (mRS 3, 4) were 55% and 41%, and favorable outcomes were achieved in 24% and 7%, respectively (detailed data are shown in **Supplementary Table 5**). Exactly 113 patients (33%) required CSF shunt surgery because of secondary hydrocephalus, and 54 (16%) were discharged with a tracheostomy.

**Figure 2.**
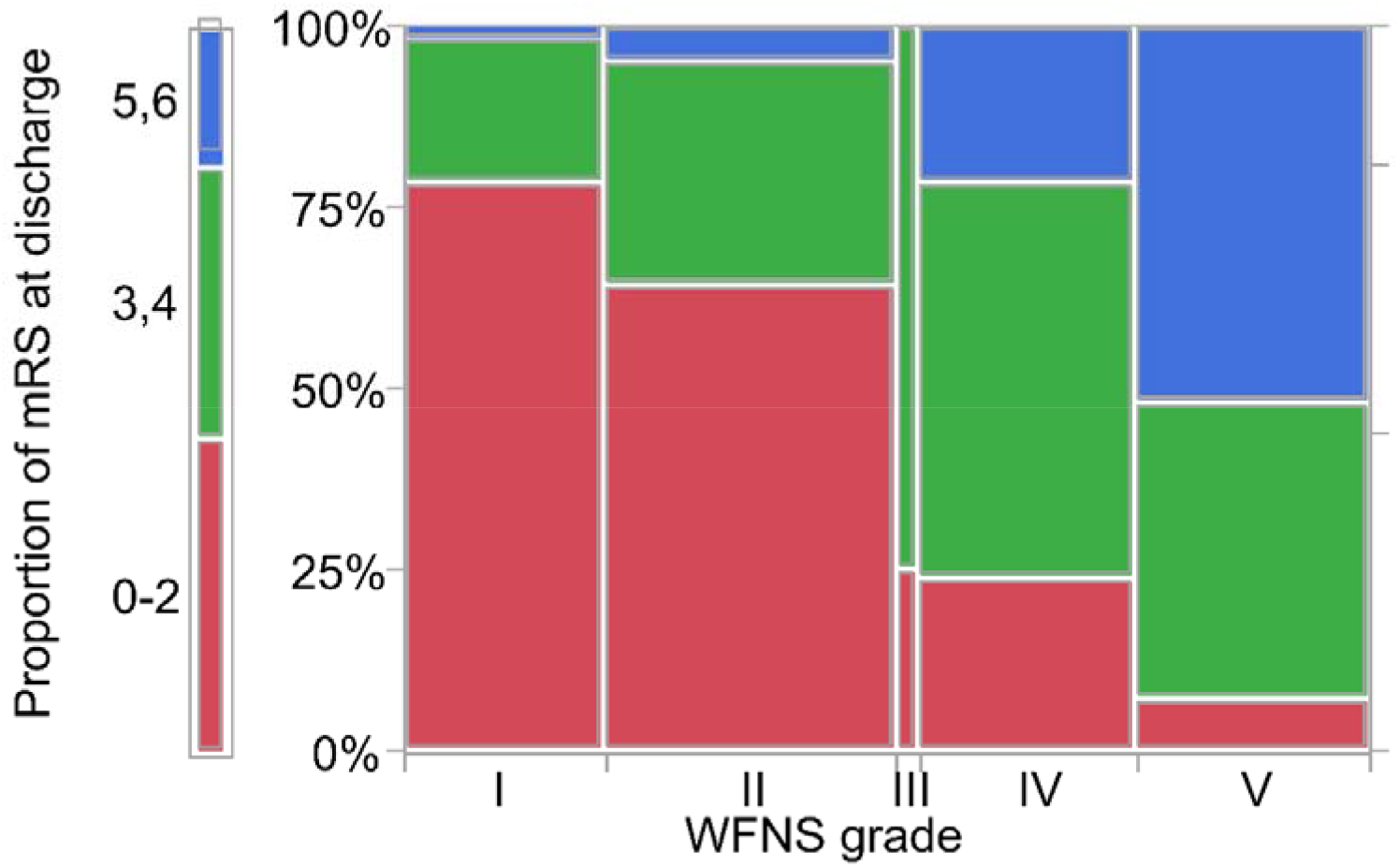
The bar graph showing association between proportion of modified Rankin scale at discharge and the initial World Federation of Neurosurgery Society grade. Modified Rankin scale was stratified into favorable (0–2), intermediate (3, 4), and severe (5, 6).

The bivariate and multivariate analysis results for factors associated with favorable outcomes are shown in **Table 5**. In the multivariate analysis, the following factors were found to be significantly negatively associated with a favorable outcome: age 65 or older (OR 0.11; 95% CI 0.06–0.22; *p* < 0.001), diabetes, anterior circulation aneurysm, concomitant with ICH, WFNS grade 4, 5, preoperative emergent VD, postoperative re-rupture, and additional postoperative surgery. Although early surgery was not associated with favorable outcomes in the bivariate analysis, this statistical significance was not observed in the multivariate analysis. The occurrence of total vasospasm or DCI did not have a statistically significant negative impact on favorable outcomes at discharge.

**TABLE 5.**
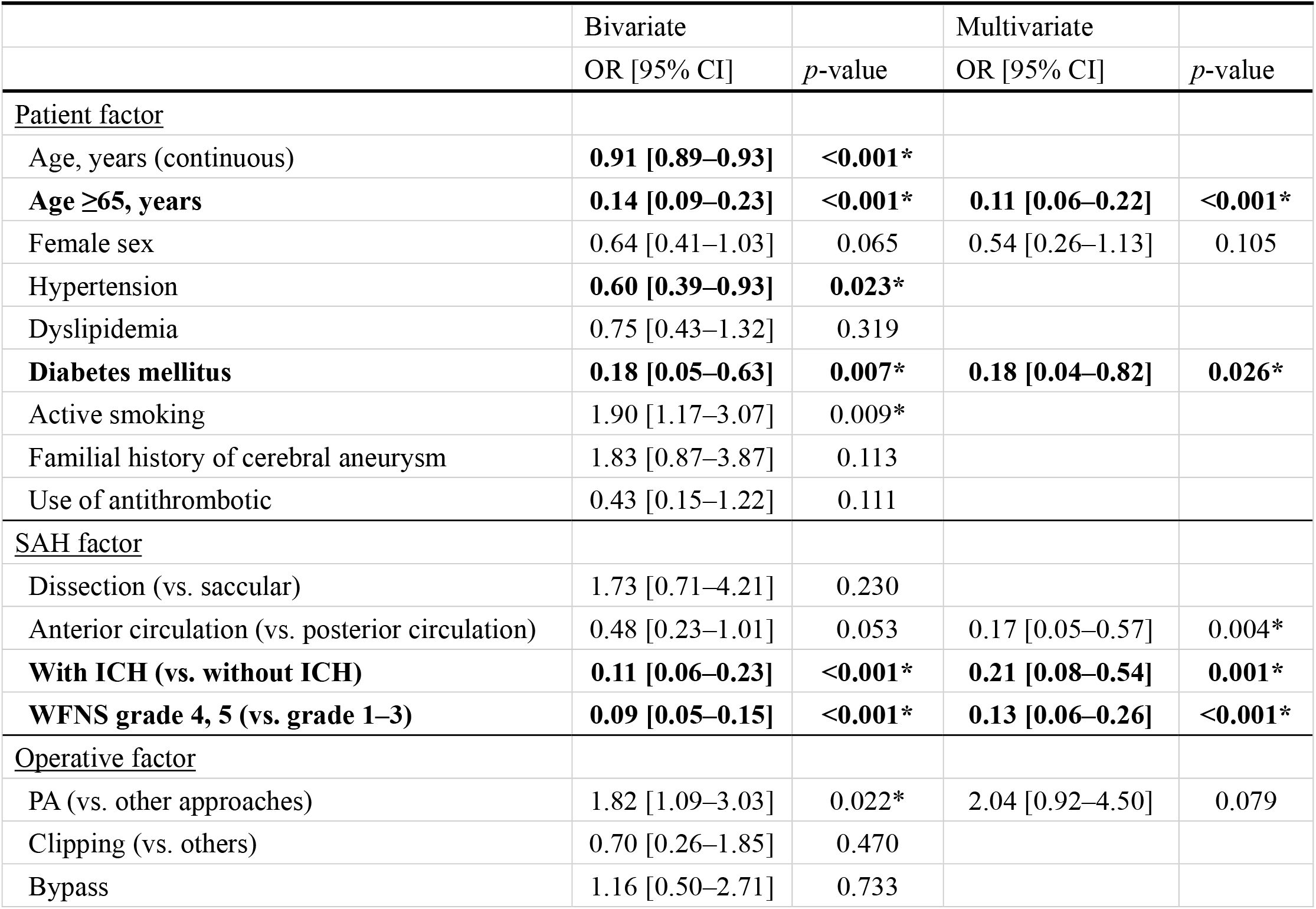
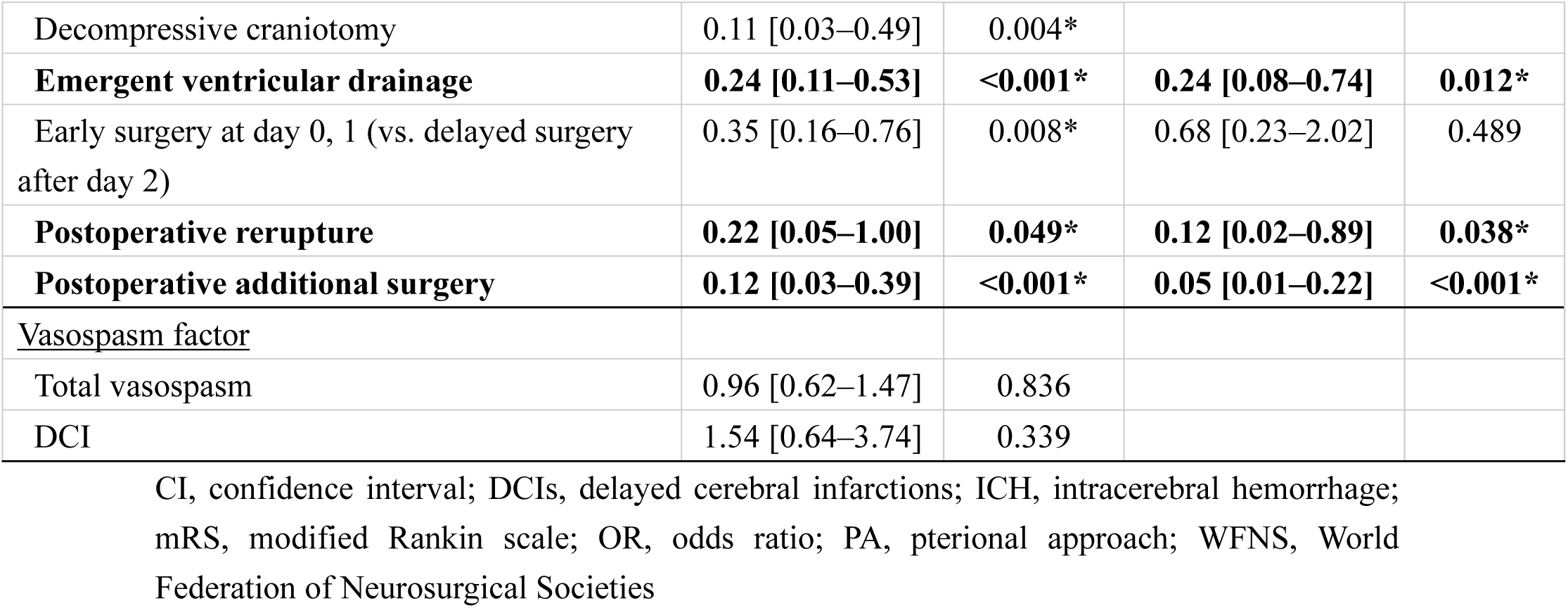
Bivariate and multivariate analyses of factors related to favorable outcomes (mRS 0–2).

|  | Bivariate |  | Multivariate |  |
| --- | --- | --- | --- | --- |
|  | OR [95% CI] | *p*-value | OR [95% CI] | *p*-value |
| **<u>Patient factor</u>** |  |  |  |  |
| Age, years (continuous) | **0.91 [0.89–0.93]** | **<0.001\*** |  |  |
| **Age ≥65, years** | **0.14 [0.09–0.23]** | **<0.001\*** | **0.11 [0.06–0.22]** | **<0.001\*** |
| Female sex | 0.64 [0.41–1.03] | 0.065 | 0.54 [0.26–1.13] | 0.105 |
| Hypertension | **0.60 [0.39–0.93]** | **0.023\*** |  |  |
| Dyslipidemia | 0.75 [0.43–1.32] | 0.319 |  |  |
| **Diabetes mellitus** | **0.18 [0.05–0.63]** | **0.007\*** | **0.18 [0.04–0.82]** | **0.026\*** |
| Active smoking | 1.90 [1.17–3.07] | 0.009* |  |  |
| Familial history of cerebral aneurysm | 1.83 [0.87–3.87] | 0.113 |  |  |
| Use of antithrombotic | 0.43 [0.15–1.22] | 0.111 |  |  |
| **<u>SAH factor</u>** |  |  |  |  |
| Dissection (vs. saccular) | 1.73 [0.71–4.21] | 0.230 |  |  |
| Anterior circulation (vs. posterior circulation) | 0.48 [0.23–1.01] | 0.053 | 0.17 [0.05–0.57] | 0.004* |
| **With ICH (vs. without ICH)** | **0.11 [0.06–0.23]** | **<0.001\*** | **0.21 [0.08–0.54]** | **0.001\*** |
| **WFNS grade 4, 5 (vs. grade 1–3)** | **0.09 [0.05–0.15]** | **<0.001\*** | **0.13 [0.06–0.26]** | **<0.001\*** |
| **<u>Operative factor</u>** |  |  |  |  |
| PA (vs. other approaches) | 1.82 [1.09–3.03] | 0.022* | 2.04 [0.92–4.50] | 0.079 |
| Clipping (vs. others) | 0.70 [0.26–1.85] | 0.470 |  |  |
| Bypass | 1.16 [0.50–2.71] | 0.733 |  |  |
| Decompressive craniotomy | 0.11 [0.03–0.49] | 0.004* |  |  |
| **Emergent ventricular drainage** | **0.24 [0.11–0.53]** | **<0.001\*** | **0.24 [0.08–0.74]** | **0.012\*** |
| Early surgery at day 0, 1 (vs. delayed surgery after day 2) | 0.35 [0.16–0.76] | 0.008* | 0.68 [0.23–2.02] | 0.489 |
| **Postoperative rerupture** | **0.22 [0.05–1.00]** | **0.049\*** | **0.12 [0.02–0.89]** | **0.038\*** |
| **Postoperative additional surgery** | **0.12 [0.03–0.39]** | **<0.001\*** | **0.05 [0.01–0.22]** | **<0.001\*** |
| <u>Vasospasm factor</u> |  |  |  |  |
| Total vasospasm | 0.96 [0.62–1.47] | 0.836 |  |  |
| DCI | 1.54 [0.64–3.74] | 0.339 |  |  |

CI, confidence interval; DCIs, delayed cerebral infarctions; ICH, intracerebral hemorrhage; mRS, modified Rankin scale; OR, odds ratio; PA, pterional approach; WFNS, World Federation of Neurosurgical Societies

## Discussion

This is the largest cohort study to report the management of vasospasms after aneurysmal SAH through VCI and continuous ICP control therapy. **Table 6** summarizes previous reports on VCI and ICP control.^13–19^ Previous studies used CD for SAH washout after surgery. VD was subsequently introduced, leading to the development of VCI. These reports indicated total vasospasm rates of 15.8%–57.8% and symptomatic vasospasm rates of 2.8%–39.5%, which align with the findings of this study.^13–19^ DCI occurrence is generally reported to be 20%–40%,^5–12^ and the reduced occurrence (0.9%–20.0%) achieved with VCI and ICP control therapy, as also found in this study, is considered favorable.^14, 15, 17, 18^ Particularly noteworthy is the excellent outcome of 0.9% for DCI achieved by Kodama et al., who used urokinase and ascorbic acid in the irrigation fluid, which requires further verification.^18^ Furthermore, systematic reviews have shown the effectiveness of intrathecal nicardipine in reducing vasospasms and DCI,^20, 21^ as well as in improving the proportion of patients with favorable outcomes (mRS 0–2).^22^ In light of the above, we explored the potential effectiveness of drainage management for ICP control to administer intrathecal nicardipine when radiological vasospasms occur. While few previous studies have described complications related to drainage management, Kodama et al. reported drain-related bleeding and ventriculitis, with a surgical requirement rate of 1.8%, but no residual sequelae.^18^ In this study, severe complications associated with drainage management were low, at 1.5%. With proficiency in the management of drainage systems, VCI and continuous ICP control can be performed safely after SAH surgery.

**Table 6.**
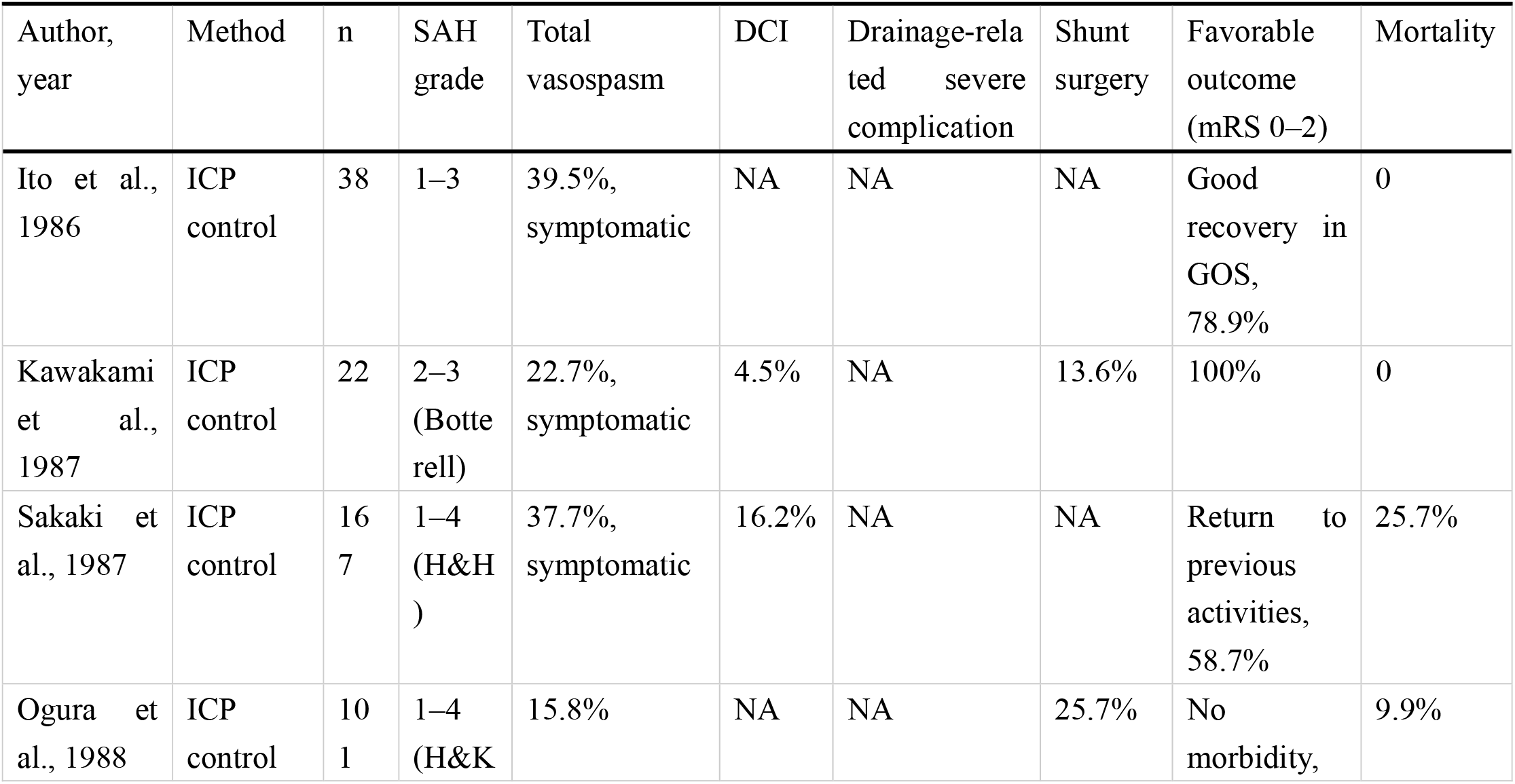
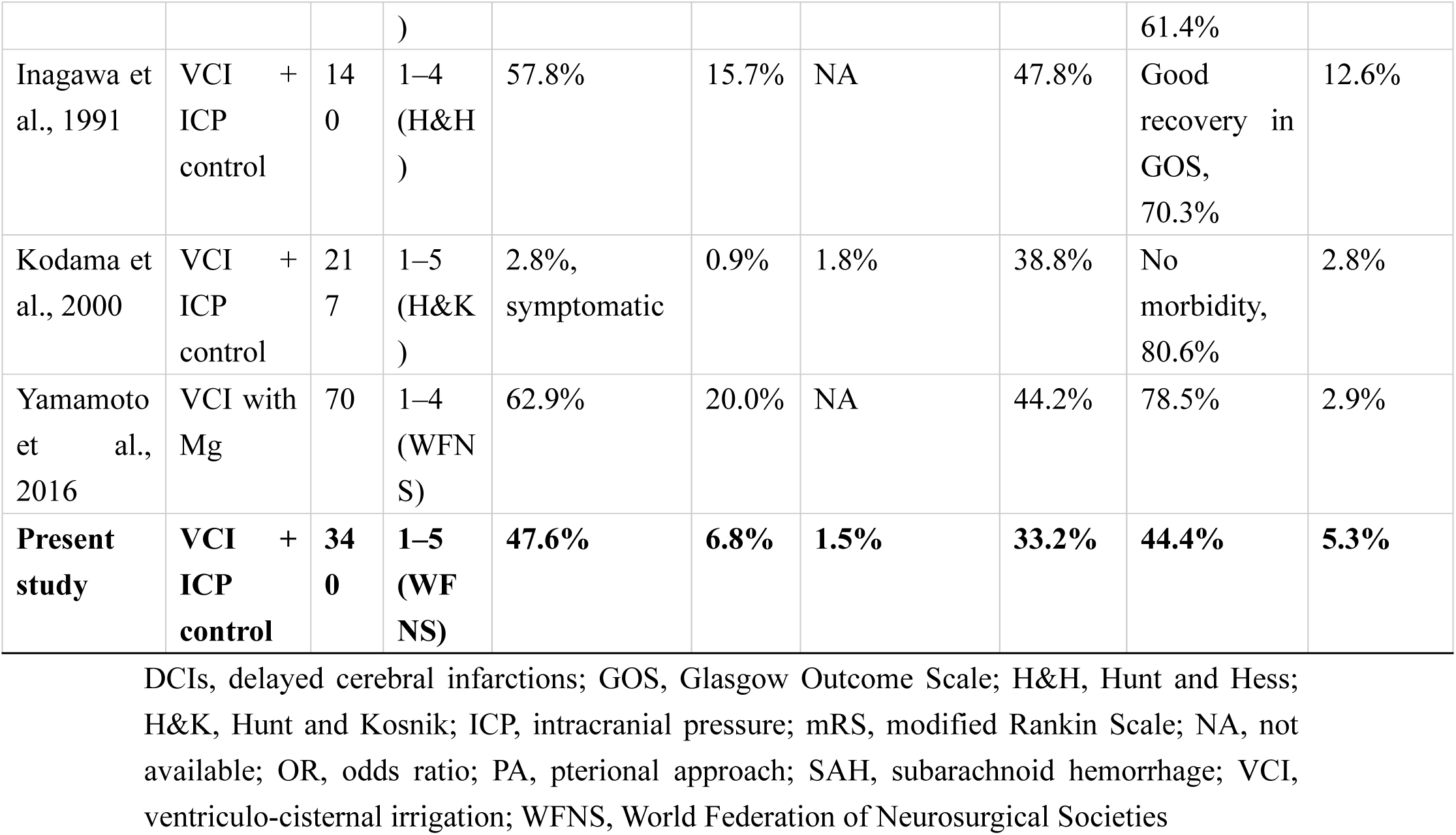
Summary of previous studies attempting to prevent cerebral vasospasm with ventriculo-cisternal irrigation and/or ICP control therapy after SAH surgery.

Early surgery was associated with a reduced risk of DCI. This result aligns with the clinical significance of VCI in removing subarachnoid clots as early as possible. Previous reports have also suggested the effectiveness of early surgery in reducing vasospasms and DCI, indicating the potential benefits of early aneurysm treatment combined with VCI and ICP control in reducing DCI.^23, 24^ Furthermore, early surgery reportedly improved outcomes in patients with SAH, and its usefulness has been incorporated into the guidelines.^2, 25–28^ Although no relationship between early surgery and favorable outcomes was observed, our findings suggest that cases requiring early surgery could be confounded by SAH severity.

This cohort included 22% of WFNS grade IV and 24% of grade V cases, indicating a population with higher severity compared to previous reports on VCI/ICP control studies, which may explain the lower proportion of favorable outcomes at 44%. While the occurrence of vasospasm or DCI was not associated with a favorable outcome in this study, the low incidence of DCI due to early surgery and management with VCI and ICP control led to the finding that DCI did not show a statistically significant positive effect on outcomes. We evaluated physical status at discharge using an assessment, potentially increasing favorable outcomes. Considering the above, compared with previous reports, favorable or intermediate outcomes, interpreted as avoiding bedridden status, were achieved in 48% of patients, even in the WFNS grade V group.^29–34^ This finding could potentially imply the effectiveness of VCI therapy in this cohort. Further studies are needed to determine whether this management approach can improve the long-term performance status of patients.

This study had several limitations. First, this was a single-center, single-arm, retrospective study without a comparative control group. Second, only patients who received VCI treatment in this study were classified as Fisher grade 2 or 3, indicating a selection bias. Thus, the analysis may have been performed in a population more prone to vasospasms. Third, the clinical treatment outcome evaluation was assessed at discharge, and not all follow-up data after discharge were available. A long-term prospective comparable study is required to evaluate the effectiveness of this treatment.

In conclusion, VCI and continuous ICP control after surgery for SAH due to a ruptured aneurysm is considered an effective and safe treatment, reducing the occurrence of DCI to 6.8%. Early surgery following the onset of SAH was particularly effective in preventing DCI in light of VCI for early cisternal clearance.

## Data Availability

The datasets generated during and/or analyzed during the current study are available from the corresponding author upon reasonable request.

## Non-standard Abbreviations and Acronyms

DCI: delayed cerebral ischemia
ICP: intracranial pressure
SAH: subarachnoid hemorrhage
VCI: ventriculo-cisternal irrigation
VD/CD: ventricular/cisternal drainage; World Federation of Neurosurgical Societies

## Acknowledgments

**None**

## Supplementary Materials

Supplementary Tables 1−5

